# Prevalence and Cumulative Incidence of Mortality Associated with Cardiovascular-Kidney-Metabolic Syndrome in the United States

**DOI:** 10.1101/2024.03.01.24303630

**Authors:** Sophie E. Claudel, Insa M. Schmidt, Sushrut S. Waikar, Ashish Verma

## Abstract

**Background:** To inform public health policies regarding disease management, it is crucial to comprehend the prevalence and mortality rates linked to various stages of Cardiovascular-Kidney-Metabolic (CKM) syndrome.

**Methods:** A longitudinal study was conducted using National Health and Nutrition Examination Survey (NHANES) data (1999-2018) from 50,624 U.S. adults, defining CKM syndrome stages based on the 2023 American Heart Association (AHA) Presidential Advisory Statement. The investigation aimed to assess changes in 10-and 15-year adjusted cumulative incidences of cardiovascular mortality with each CKD syndrome stage and describe the cross-sectional prevalence of CKM syndrome from 1999 to 2020.

**Results:** CKM syndrome prevalence by stage was as follows: Stage 0, 12.5% (95% CI, 12.0-12.9); Stage 1, 16.7% (95% CI, 16.2-17.2); Stage 2, 40.0% (95% CI, 38.4-39.6); Stage 3, 22.9% (95% CI, 22.5-23.4); Stage 4, 8.9% (95% CI, 8.6-9.2). Over a median 9.5-year follow-up, 2,557 participants experienced cardiovascular death. The proportion of participants in Stages 0 and 3 decreased, while Stage 1 increased. The 15-year adjusted cumulative incidences of cardiovascular mortality were: Stage 0, 4.8% (95% CI 3.1-6.6); Stage 1, 5.3% (95% CI 4.0-6.6); Stage 2, 7.9% (95% CI 8.1-10.3); Stage 3, 9.2% (95% CI 8.1-10.3); Stage 4, 15.6% (95% CI 14.7-16.6). The absolute risk difference between CKM Stage 4 and Stage 0 at 15 years was 10.8% (95% CI 8.8-12.8).

**Conclusions:** Our findings showed a graded increase in cardiovascular mortality associated with each CKM stage. The trends observed by stage prevalence emphasize critical opportunities for stabilizing risk factors, thereby preventing adverse cardiovascular outcomes.

## Introduction

In October 2023, the American Heart Association (AHA) released a Presidential Advisory on Cardiovascular-Kidney-Metabolic (CKM) syndrome. CKM syndrome is a clinical framework and disease staging system that describes the intersection of diabetes, obesity, chronic kidney disease, and cardiovascular disease.^1,2^ There are five stages of CKM syndrome, ranging from a state of health (Stage 0) to clinical cardiovascular disease (Stage 4). Early CKM syndrome is thought to develop due to excess or dysfunctional adiposity.^2,3^ As the prevalence of overweight and obesity is steadily rising in the United States (U.S.), CKM syndrome may pose an increasing threat to public health.^4^ However, the absolute risk or mortality associated with each stage of CKM syndrome must be better understood. Examining the current scope of CKM syndrome in the U.S. and characterizing the relationship between CKM syndrome and mortality are fundamental to understanding the public health implications of the newly defined framework.

The objectives of this study were to summarize the prevalence and trends of CKM syndrome in the U.S. over 20 years and to estimate the adjusted cumulative incidence of cardiovascular mortality associated with each CKM stage using data from the U.S. National Health and Nutrition Examination Survey (NHANES).

## Methods

### Study Population

We included adults aged ≥20 years who participated in NHANES between 1999-2020. NHANES is a biannual, cross-sectional survey of the non-institutionalized U.S. population that provides demographic, social, and medical surveys, in addition to laboratory and examination data.^5^ We excluded participants who were pregnant at the time of the exam (N=1324), had undergone dialysis in the prior 12 months (N=82), or were missing mortality data (N=83). We used the 1999-2020 survey years for prevalence estimates (N=53,959) and the 1999-2018 survey years for the analyses assessing mortality (N=50,624), as linkage to the U.S. National Death Index was only available through December 31^st^, 2019.

### Primary Exposure

Primary exposure was CKM stages as a categorical variable. We defined the stages of CKM syndrome per the American Heart Association (AHA) Presidential Advisory Statement on CKM syndrome, as outlined in **Table S1**.^1.^ The stages are as follows: Stage 0, no CKM risk factors; Stage 1, excess or dysfunctional adiposity; Stage 2, metabolic risk factors or chronic kidney disease; Stage 3, subclinical cardiovascular disease; and Stage 4, clinical cardiovascular disease. As recommended by the AHA, we incorporated the novel 10-year PREVENT (Predicting Risk of Cardiovascular Disease Events) risk equation.^6,7^

### Ascertainment of Covariates

Data obtained at the baseline survey years included sociodemographic characteristics, detailed medical history, comprehensive medication lists, standardized blood pressure measurements, anthropometric measures, and laboratory measurements. Detailed measurement and laboratory methods for all NHANES variables can be found online.^5^ The eGFR was calculated using the 2021 CKD-EPI_cr_ equation.^9^

### Missing values

Several covariates had missing values (**Table S2**) and were assumed to be missing at random. We imputed systolic blood pressure, serum creatinine, waist circumference, BMI, hemoglobin A1c, and UACR with hot deck multiple imputation using simple random sampling with replacement and accounting for complex survey design.^10^ The sample characteristics before imputation are shown in **Table S3** for comparison.

### Ascertainment of mortality

For analyses assessing mortality, we included NHANES years 1999-2018. We ascertained mortality through linkage to the U.S. National Death Index through December 31^st^, 2019. We defined cardiovascular mortality as death from diseases of the heart (International Classification of Disease (ICD)-10 codes I00-I09, I11, I13, I20-I151) and cerebrovascular diseases (ICD-10 codes I60-I69).

### Statistical Analysis

We calculated the weighted, age-adjusted prevalence of each CKM syndrome stage. Estimates were age-standardized to the 2010 U.S. Census. To minimize the effect of small sample sizes, we created five pre-determined periods for analysis: 1999-2002, 2003-2006, 2007-2010, 2011-2014, and 2015-2020. The 2019-2020 NHANES survey cycle was prematurely stopped in March 2020 due to the COVID-19 pandemic. Therefore, NHANES provides a 2017-2020 combined file that is nationally representative of the U.S. population despite early termination of the survey, and which may be used for prevalence estimates.^11^ We calculated prevalence estimates and examined trends in prevalence for the overall sample by age group (20-39 years, 40-59 years, and ≥60 years) and by race or ethnicity. As ‘non-Hispanic Asian’ was first included as a separate race category in 2011, we provide biannual estimates of CKM syndrome prevalence for non-Hispanic Asians from 2011-2020 only. We used standard Wald tests from survey logistic regression models to obtain *p*-values for temporal trends, controlling for age.^12^

We then calculated the 10- and 15-year adjusted cumulative incidences and cumulative incidence functions using confounder-adjusted survival curves employing the G-Formula method for covariate adjustment.^13^ We plotted adjusted cumulative incidence and cumulative incidence function curves using the R software package “adjustedCurves” using final multivariable-adjusted cause-specific Cox proportional hazard and Fine and Gray competing risk models, respectively.^13^ The competing risk regression was done using R software’s “cmprsk” package. We confirmed no violations of the proportional hazard’s assumption by Schoenfeld residuals using the “cox. zph” function in the “survival” R package.^14^ We calculated the absolute risk differences between Stage 4 and each earlier stage of CKM syndrome and the corresponding 95% confidence interval (CI) at 10 and 15 years of follow-up. The adjustment strategy was based on covariates’ biological and clinical plausibility as potential confounders of the association between CKM syndrome and cardiovascular mortality. Models were adjusted for age, sex, self-reported race or ethnicity (non-Hispanic White, non-Hispanic Black, Mexican American, other Hispanic, and other race), survey year, body mass index (BMI, kg/m^2^), waist circumference (cm), total cholesterol (mg/dL), use of a statin, use of an angiotensin-converting enzyme inhibitor (ACEi) or angiotensin receptor blocker (ARB), systolic blood pressure (mmHg), smoking status (never/former vs. current), hemoglobin A1c (%), urinary albumin-to-creatinine ratio (UACR) (mg/g), and estimated glomerular filtration rate (eGFR) (ml/min per 1.73m^2^), history of chronic respiratory disease, history of cancer, and history of liver disease. In a sensitivity analysis, we additionally adjusted for the following social determinants of health: education, health insurance, food security, and poverty-to-income ratio. We then performed a sensitivity analysis, repeating the analysis using all-cause mortality as the outcome.

We additionally describe the relative risk of mortality using multivariable-adjusted Cox regression models. The proportional hazards assumption was tested with Schoenfeld residuals. The adjustment strategy was the same as described for the cumulative incidence curves. We performed sensitivity analyses using both all-cause and non-cardiovascular mortality as the outcome and also adjusting for social determinants of health. The social determinants of health included educational attainment, health insurance, food security, and poverty-to-income ratio.

The adjusted cumulative incidence curves were unweighted as no established procedure exists to incorporate survey weights into adjusted cumulative incidence models. All other analyses accounted for the complex survey design of NHANES. As the adjusted cumulative incidence curves were unweighted, we additionally provide unweighted hazard ratios to compare to the primary analysis.

Analyses were performed in SAS (version 9.4) or R software (version 3.5). Data were analyzed by S.E.C and A.V. A two-tailed p-value <0.05 was considered statistically significant. The width of the confidence intervals was not adjusted for multiplicity. All data are publicly available at https://wwwn.cdc.gov/nchs/nhanes/default.aspx.

## Results

The characteristics of the study population are shown by CKM syndrome stage in **Table 1**. The study population mean (standard deviation) age was 47.3 (17.0) years, and 51.3% were women. The median length of follow-up was 9.5 years [interquartile range 4.9 to 14.5], and there were 2,557 cardiovascular deaths (8,169 all-cause mortality events).

**Table 1.**
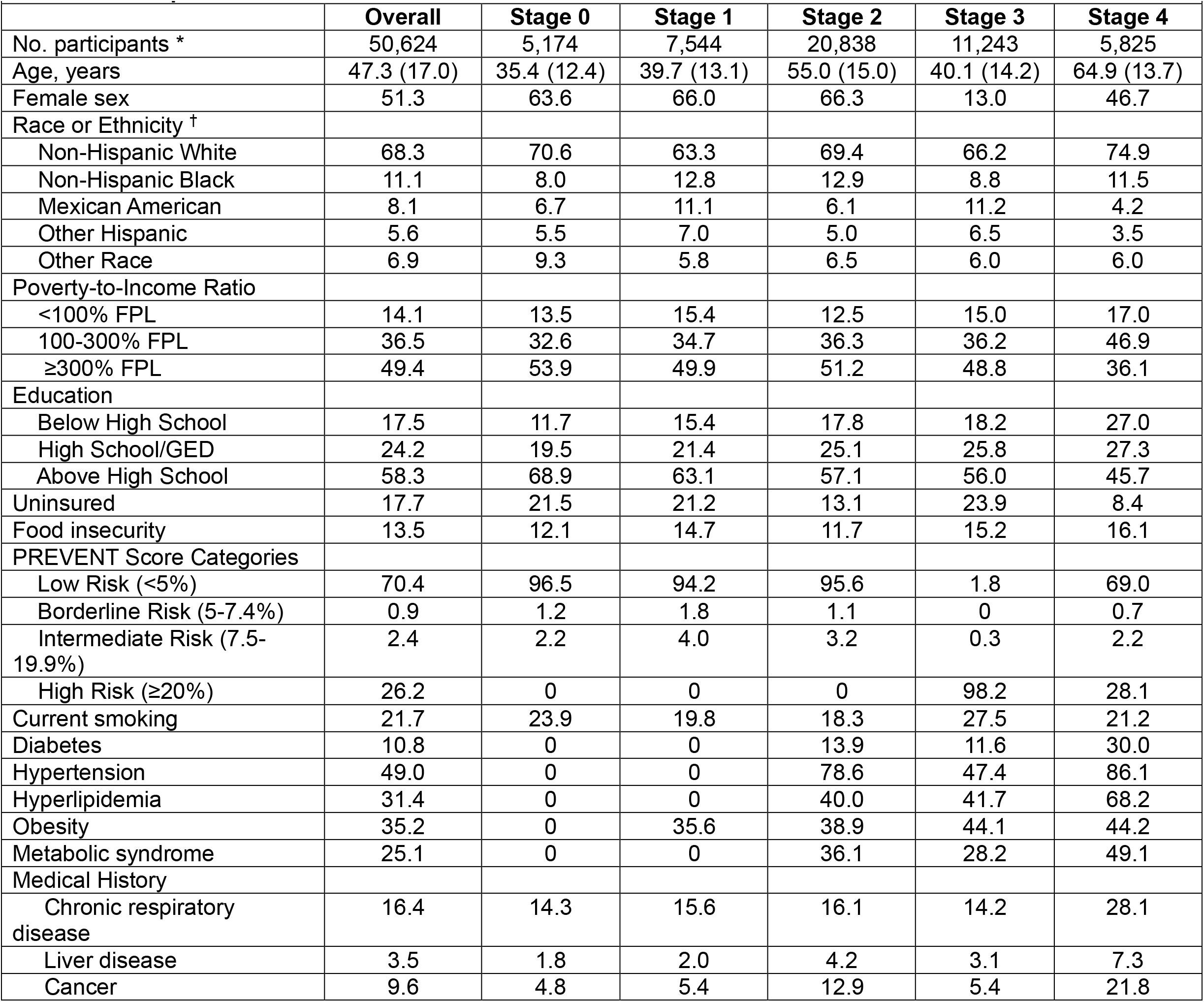
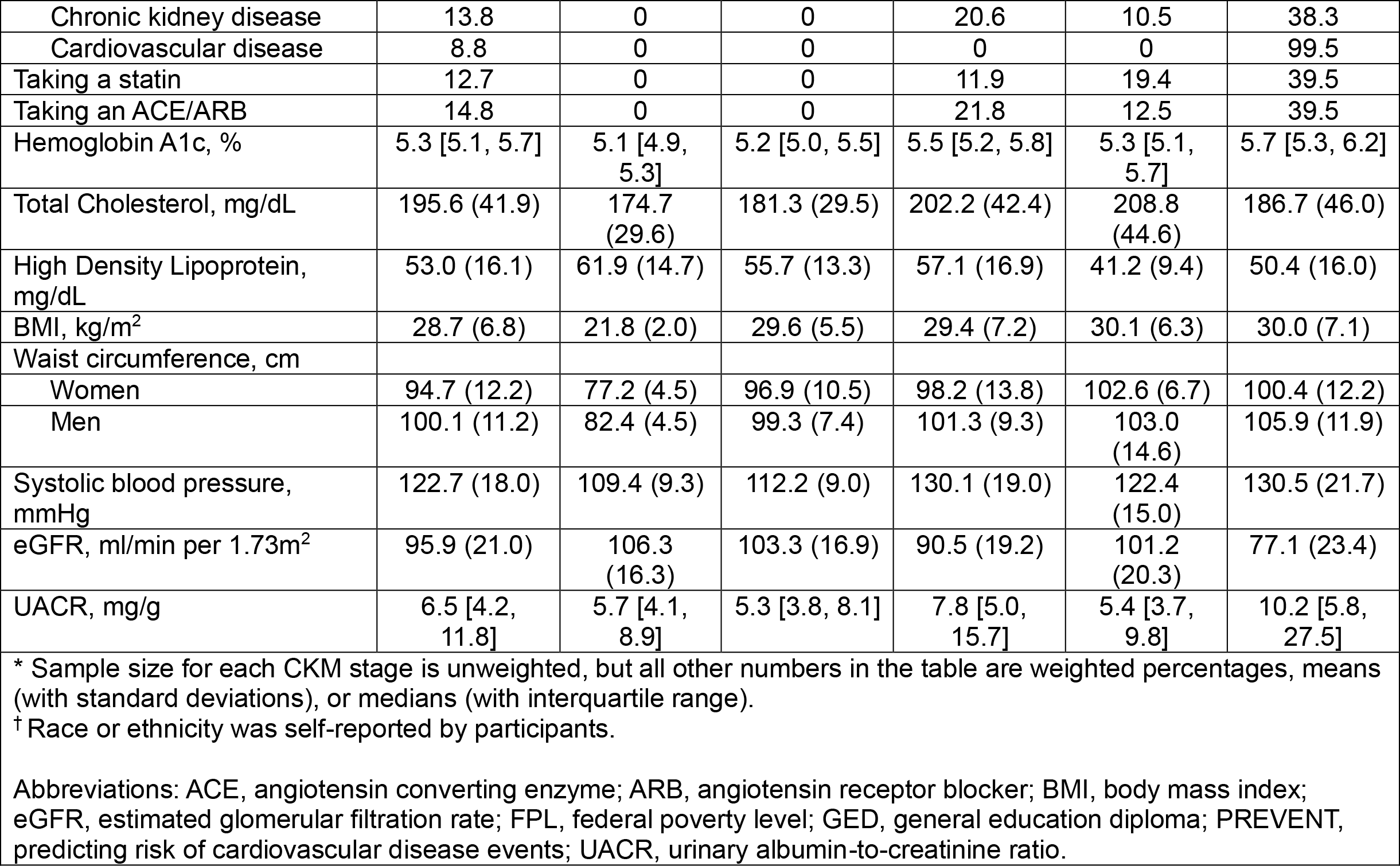
Participant Characteristics.

| **Table 1. Participant Characteristics** |  |  |  |  |  |  |
| --- | --- | --- | --- | --- | --- | --- |
|  | **Overall** | **Stage 0** | **Stage 1** | **Stage 2** | **Stage 3** | **Stage 4** |
| No. participants * | 50,624 | 5,174 | 7,544 | 20,838 | 11,243 | 5,825 |
| Age, years | 47.3 (17.0) | 35.4 (12.4) | 39.7 (13.1) | 55.0 (15.0) | 40.1 (14.2) | 64.9 (13.7) |
| Female sex | 51.3 | 63.6 | 66.0 | 66.3 | 13.0 | 46.7 |
| Race or Ethnicity <sup>†</sup> |  |  |  |  |  |  |
| Non-Hispanic White | 68.3 | 70.6 | 63.3 | 69.4 | 66.2 | 74.9 |
| Non-Hispanic Black | 11.1 | 8.0 | 12.8 | 12.9 | 8.8 | 11.5 |
| Mexican American | 8.1 | 6.7 | 11.1 | 6.1 | 11.2 | 4.2 |
| Other Hispanic | 5.6 | 5.5 | 7.0 | 5.0 | 6.5 | 3.5 |
| Other Race | 6.9 | 9.3 | 5.8 | 6.5 | 6.0 | 6.0 |
| Poverty-to-Income Ratio |  |  |  |  |  |  |
| <100% FPL | 14.1 | 13.5 | 15.4 | 12.5 | 15.0 | 17.0 |
| 100-300% FPL | 36.5 | 32.6 | 34.7 | 36.3 | 36.2 | 46.9 |
| ≥300% FPL | 49.4 | 53.9 | 49.9 | 51.2 | 48.8 | 36.1 |
| Education |  |  |  |  |  |  |
| Below High School | 17.5 | 11.7 | 15.4 | 17.8 | 18.2 | 27.0 |
| High School/GED | 24.2 | 19.5 | 21.4 | 25.1 | 25.8 | 27.3 |
| Above High School | 58.3 | 68.9 | 63.1 | 57.1 | 56.0 | 45.7 |
| Uninsured | 17.7 | 21.5 | 21.2 | 13.1 | 23.9 | 8.4 |
| Food insecurity | 13.5 | 12.1 | 14.7 | 11.7 | 15.2 | 16.1 |
| PREVENT Score Categories |  |  |  |  |  |  |
| Low Risk (<5%) | 70.4 | 96.5 | 94.2 | 95.6 | 1.8 | 69.0 |
| Borderline Risk (5-7.4%) | 0.9 | 1.2 | 1.8 | 1.1 | 0 | 0.7 |
| Intermediate Risk (7.5-19.9%) | 2.4 | 2.2 | 4.0 | 3.2 | 0.3 | 2.2 |
| High Risk (≥20%) | 26.2 | 0 | 0 | 0 | 98.2 | 28.1 |
| Current smoking | 21.7 | 23.9 | 19.8 | 18.3 | 27.5 | 21.2 |
| Diabetes | 10.8 | 0 | 0 | 13.9 | 11.6 | 30.0 |
| Hypertension | 49.0 | 0 | 0 | 78.6 | 47.4 | 86.1 |
| Hyperlipidemia | 31.4 | 0 | 0 | 40.0 | 41.7 | 68.2 |
| Obesity | 35.2 | 0 | 35.6 | 38.9 | 44.1 | 44.2 |
| Metabolic syndrome | 25.1 | 0 | 0 | 36.1 | 28.2 | 49.1 |
| Medical History |  |  |  |  |  |  |
| Chronic respiratory disease | 16.4 | 14.3 | 15.6 | 16.1 | 14.2 | 28.1 |
| Liver disease | 3.5 | 1.8 | 2.0 | 4.2 | 3.1 | 7.3 |
| Cancer | 9.6 | 4.8 | 5.4 | 12.9 | 5.4 | 21.8 |
| Chronic kidney disease | 13.8 | 0 | 0 | 20.6 | 10.5 | 38.3 |
| Cardiovascular disease | 8.8 | 0 | 0 | 0 | 0 | 99.5 |
| Taking a statin | 12.7 | 0 | 0 | 11.9 | 19.4 | 39.5 |
| Taking an ACE/ARB | 14.8 | 0 | 0 | 21.8 | 12.5 | 39.5 |
| Hemoglobin A1c, % | 5.3 [5.1, 5.7] | 5.1 [4.9, 5.3] | 5.2 [5.0, 5.5] | 5.5 [5.2, 5.8] | 5.3 [5.1, 5.7] | 5.7 [5.3, 6.2] |
| Total Cholesterol, mg/dL | 195.6 (41.9) | 174.7 (29.6) | 181.3 (29.5) | 202.2 (42.4) | 208.8 (44.6) | 186.7 (46.0) |
| High Density Lipoprotein, mg/dL | 53.0 (16.1) | 61.9 (14.7) | 55.7 (13.3) | 57.1 (16.9) | 41.2 (9.4) | 50.4 (16.0) |
| BMI, kg/m<sup>2</sup> | 28.7 (6.8) | 21.8 (2.0) | 29.6 (5.5) | 29.4 (7.2) | 30.1 (6.3) | 30.0 (7.1) |
| Waist circumference, cm |  |  |  |  |  |  |
| Women | 94.7 (12.2) | 77.2 (4.5) | 96.9 (10.5) | 98.2 (13.8) | 102.6 (6.7) | 100.4 (12.2) |
| Men | 100.1 (11.2) | 82.4 (4.5) | 99.3 (7.4) | 101.3 (9.3) | 103.0 (14.6) | 105.9 (11.9) |
| Systolic blood pressure, mmHg | 122.7 (18.0) | 109.4 (9.3) | 112.2 (9.0) | 130.1 (19.0) | 122.4 (15.0) | 130.5 (21.7) |
| eGFR, ml/min per 1.73m<sup>2</sup> | 95.9 (21.0) | 106.3 (16.3) | 103.3 (16.9) | 90.5 (19.2) | 101.2 (20.3) | 77.1 (23.4) |
| UACR, mg/g | 6.5 [4.2, 11.8] | 5.7 [4.1, 8.9] | 5.3 [3.8, 8.1] | 7.8 [5.0, 15.7] | 5.4 [3.7, 9.8] | 10.2 [5.8, 27.5] |

\* Sample size for each CKM stage is unweighted, but all other numbers in the table are weighted percentages, means (with standard deviations), or medians (with interquartile range).
† Race or ethnicity was self-reported by participants.
Abbreviations: ACE, angiotensin converting enzyme; ARB, angiotensin receptor blocker; BMI, body mass index; eGFR, estimated glomerular filtration rate; FPL, federal poverty level; GED, general education diploma; PREVENT, predicting risk of cardiovascular disease events; UACR, urinary albumin-to-creatinine ratio.

### Prevalence of CKM syndrome by stage

The overall prevalence of CKM syndrome by stage was as follows: Stage 0, 12.5% (95% CI, 12.0 to 12.9); Stage 1, 16.7% (95% CI, 16.2 to 17.2); Stage 2, 40.0% (95% CI, 38.4 to 39.6); Stage 3, 22.9% (95% CI, 22.5 to 23.4); Stage 4, 8.9% (95% CI, 8.6 to 9.2) (**Figure 1, Table S4**).

**Figure 1.**
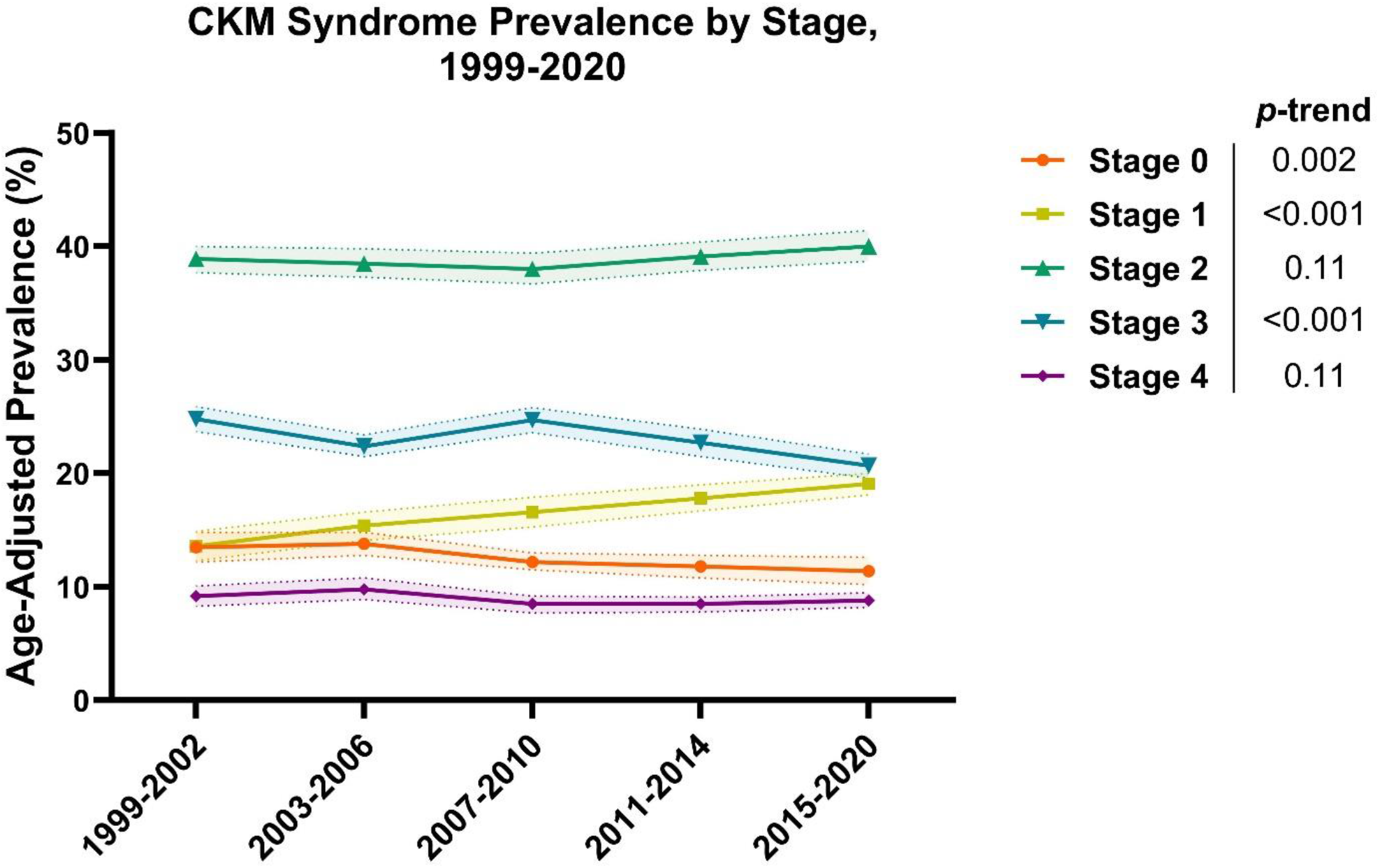
Age-adjusted prevalence of CKM syndrome by stage, 1999-2020. Prevalence estimates were age-standardized to the 2010 U.S. Census. *p*-trend was significant for CKM Stage 0, Stage 1, and Stage 3.

The proportion of patients with CKM syndrome Stage 0 decreased from 13.5% (95% CI, 12.2 to 14.8) in 1999-2002 to 11.4% (95% CI, 10.2 to 12.6) in 2015-2020, while the proportion of patients with Stage 1 increased from 13.6% (95% CI, 12.3 to 14.9) to 19.1% (95% CI, 18.1 to 20.0) over the same period.

### Prevalence of CKM syndrome stages by demographic characteristics

The prevalence of Stage 2 CKM syndrome was highest among women, whereas Stage 3 was most prevalent among men (**Table S5**). Among adults ages 40-59 years, the prevalence of Stage 0 CKM syndrome decreased from 11.8% (95% CI, 9.9 to 13.7) to 7.6% (95% CI, 6.3 to 8.9), while the prevalence of Stage 1 CKM syndrome increased from 14.4% (95% CI, 12.5 to 16.2) to 19.3% (95% CI, 17.6 to 21.1) (**Table S6**). The prevalence of Stage 3 CKM syndrome increased among older adults (age ≥60 years) and decreased among adults aged 20-59 years. Young adults demonstrated an increase in Stage 1 CKM syndrome from 19.1% (95% CI, 17.2 to 20.8) to 27.6% (95% CI, 25.8 to 29.4) between 1999-2020 (**Table S6**). A more significant proportion of non-Hispanic White and non-Hispanic Asian participants were in CKM Stage 0, and a more substantial proportion of non-Hispanic Black participants were in CKM Stages 2 and 4 compared to other racial and ethnic groups (**Tables S7**). Trends in prevalence by race or ethnicity are described in **Tables S8-9**.

### Association between CKM syndrome stages and cardiovascular mortality

The 15-year adjusted cumulative incidences of cardiovascular mortality associated with CKM syndrome were as follows: Stage 0, 4.8% (95% CI, 3.1 to 6.6); Stage 1, 5.3% (95% CI, 4.0 to 6.6); Stage 2, 7.9% (95% CI, 7.4 to 8.5); Stage 3, 9.2% (95% CI 8.1 to 10.3); Stage 4, 15.6% (95% CI, 14.7 to 16.6) (**Figure 1**). The absolute risk difference between Stage 4 and Stage 0,

Stage 1, Stage 2, and Stage 3 were 10.8% (95% CI, 8.8 to 12.8), 10.3% (95% CI, 8.7 to 11.9), 7.6% (95% CI, 6.6 to 8.7), and 6.4% (95% CI, 5.0 to 7.5), respectively.

The graded risk across increasing CKM syndrome stages was qualitatively similar when examining the adjusted cumulative incidence function for cardiovascular mortality (**Figure 2**). The 15-year adjusted cumulative incidence functions for cardiovascular mortality at each stage were as follows: Stage 0, 3.4% (95% CI, 2.1 to 4.7); Stage 1, 3.9% (95% CI, 2.9 to 4.8); Stage 2, 5.7% (95% CI, 5.3 to 6.1); Stage 3, 6.2% (95% CI, 5.4 to 6.9); Stage 4, 10.9% (95% CI, 10.1 to 11.6). The absolute risk differences for the cumulative incidence function are shown in **Figure 2**.

**Figure 2.**
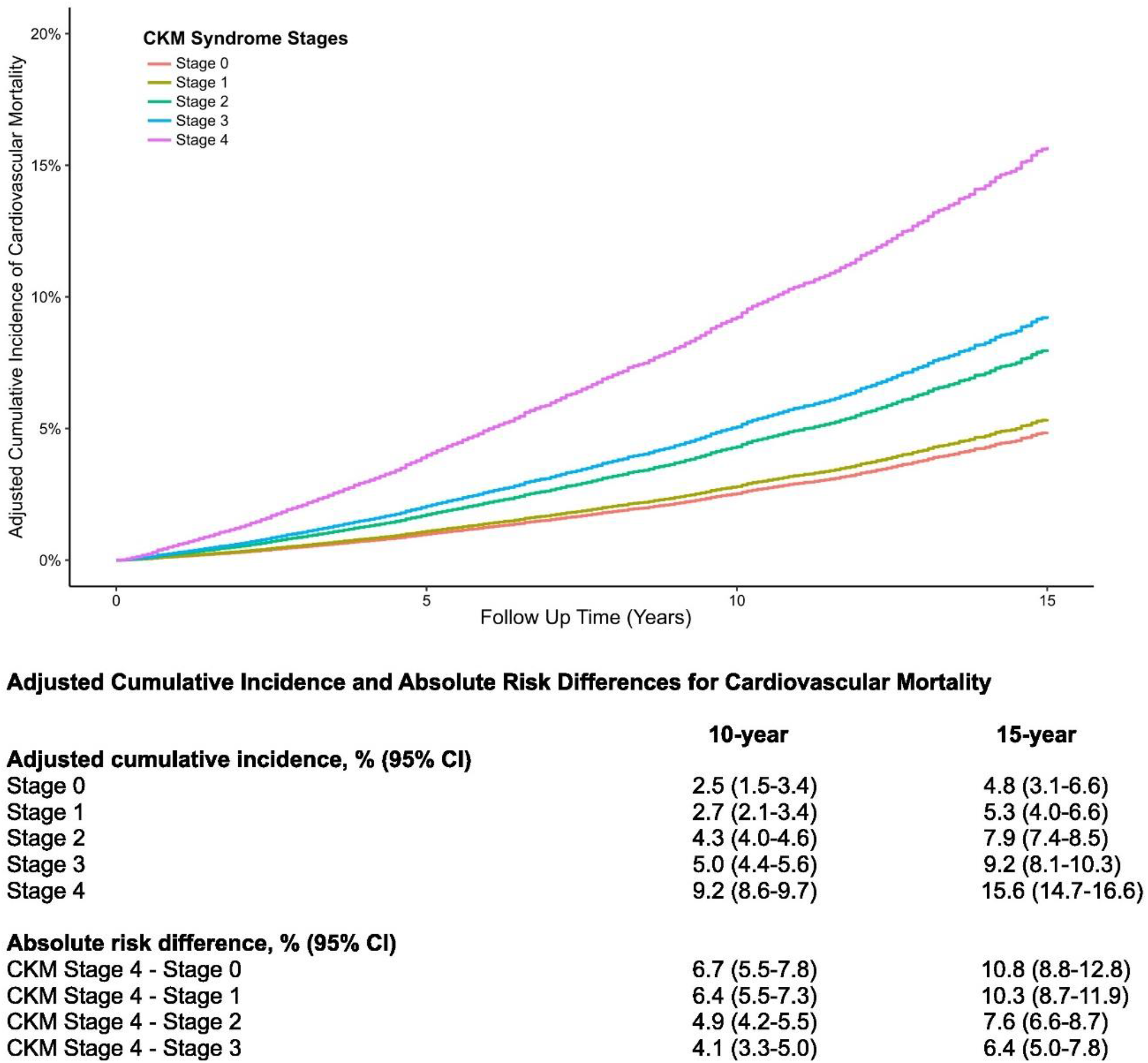
Adjusted cumulative incidences and 10-and 15-year risk differences in adjusted cumulative incidences of cardiovascular mortality by CKM stage. The adjusted cumulative incidences were calculated using confounder-adjusted survival curves employing the G-Formula method for covariate adjustment. Survey weights were not used in calculating cumulative incidence.

**Figure 3.**
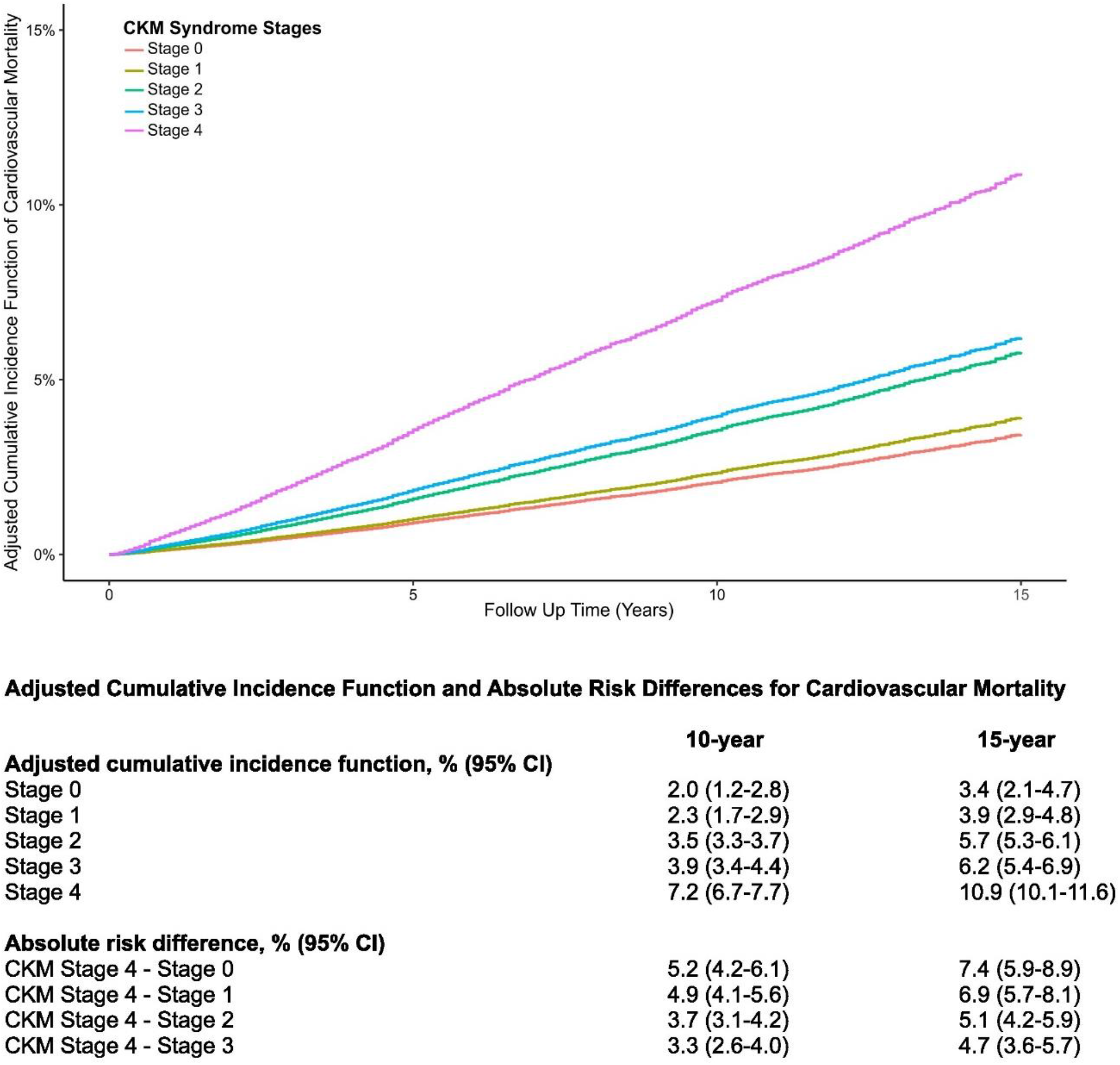
Adjusted cumulative incidences and 10-and 15-year risk differences in adjusted cumulative incidences of cardiovascular mortality by CKM stage. The adjusted cumulative incidence functions (derived from competing risk models) were calculated using confounder-adjusted survival curves employing the G-Formula method for covariate adjustment. Survey weights were not used in calculating cumulative incidence functions.

Compared to CKM syndrome Stage 0, Stage 4 was associated with 3.93-fold [95% CI, 2.30 to 6.70] higher cardiovascular mortality (**Table 2**). The association was graded by CKM syndrome stage, where the hazard ratios for cardiovascular mortality were 1.90 [95% CI, 1.10 to 3.25], 1.60 [95% CI, 0.96 to 2.67], and 1.06 [95% CI 0.63 to 1.79] for Stages 3, 2, and 1, respectively. Adjusting for social determinants of health modestly attenuated the hazard ratios (**Table S10**). **Table S11** demonstrates unweighted Cox proportional hazards regression models.

**Table 2.** Association between CKM syndrome stage and cardiovascular mortality.

|  | **Stage 0** | **Stage 1** | **Stage 2** | **Stage 3** | **Stage 4** |
| --- | --- | --- | --- | --- | --- |
| No. events | 27 | 60 | 1,143 | 256 | 1,071 |
| No. participants | 5,174 | 7,544 | 20,838 | 11,243 | 5,825 |
| Crude | Referent | 1.46 [0.87, 2.46] | 10.27 [6.43, 16.40] | 3.73 [2.29, 6.08] | 54.34 [33.89, 87.10] |
| Multivariable model 1<sup>a</sup> | Referent | 0.97 [0.58, 1.62] | 1.77 [1.11, 2.83] | 2.05 [1.24, 3.38] | 4.76 [2.93, 7.74] |
| Multivariable model 2<sup>b</sup> | Referent | 0.97 [0.58, 1.63] | 1.49 [0.91, 2.46] | 1.77 [1.05, 3.01] | 3.72 [2.21, 6.26] |
| Multivariable model 3<sup>c</sup> | Referent | 1.06 [0.63, 1.79] | 1.60 [0.96, 2.67] | 1.90 [1.10, 3.25] | 3.93 [2.30, 6.70] |

<sup>a</sup> Multivariable model 1: age, sex, race or ethnicity, smoking status, survey year.
<sup>b</sup> Multivariable model 2: adjusted for Model 2 + BMI, waist circumference, hemoglobin A1c, systolic blood pressure, total cholesterol, statin use, ACE/ARB use, UACR, eGFR.
<sup>c</sup> Multivariable model 3: adjusted for Model 3 + history of chronic respiratory disease, history of liver disease, history of cancer.

### Association between CKM syndrome stages and all-cause and non-cardiovascular mortality

Results were qualitatively similar when examining the relationship between CKM syndrome stages and all-cause mortality, except for a marginally higher cumulative incidence of mortality with Stage 0 compared to Stage 1 CKM syndrome (**Table S12, Figure S1**). In proportional hazards models, the multivariable-adjusted hazard ratio for all-cause mortality was 2.06 [95% CI, 1.72 to 2.48] for Stage 4 CKM syndrome (**Table S13**). Adjusting for social determinants of health attenuated the hazard ratio to 1.87 [95% CI, 1.53 to 2.27] (**Table S10**). There was also a graded, but attenuated, association between CKM syndrome stages and non-cardiovascular mortality (**Table S14**).

## Discussion

This is the first report on the prevalence of CKM syndrome in the U.S. and describes the graded risk of cardiovascular mortality associated with each CKM stage. The staging system was developed based on accumulated clinical and pathophysiologic data and our findings demonstrate that the stages correlate well with cumulative risk for cardiovascular death.^2^ Furthermore, the associations between CKM syndrome stage and cardiovascular mortality remained robust in competing risk analyses, a fundamental knowledge gap highlighted by Ndumele *et al*,^2^ and when adjusting for social determinants of health. This knowledge adds to clinicians’ armamentarium for counseling patients on cardiovascular risk by providing absolute risk estimates associated with each stage. Simultaneously, our prevalence data establish population-level metrics by which public health practitioners and health policy experts can measure the impact of future interventions.

Between 1999 and 2020, we observed an increase in Stage 1 CKM syndrome across all sociodemographic subgroups, frequently corresponding with a decrease in Stage 0. This suggests an increasing population at risk for progressive CKM syndrome. Improving access to obesity treatments – pharmacological, surgical, and behavioral – may lead to stabilization of the population in Stage 1 and thereby prevent clinical complications of dysfunctional adiposity.^15–17^ Early and aggressive treatment of obesity may even lead to regression of CKM syndrome in specific individuals, as weight loss can lead to improved insulin sensitivity.^18–20^ Stage 1 CKM syndrome should not be the sole focus of preventative measures. Our data also suggest opportunities exist to prevent progression even in Stage 3, as evidenced by the 6.4% risk difference in cardiovascular mortality between Stage 3 and Stage 4 and low rates of cardioprotective medication use among these participants in our population and among prior studies.^21–24^ Additionally, the prevalence of well-controlled diabetes and hypertension has declined among NHANES participants in recent years.^8,25^ This suggests that individuals in Stages 2 and 3 are at high risk of CKM syndrome progression and adverse cardiovascular outcomes. Appropriate application of known kidney-cardiovascular therapies, including sodium-glucose co-transporter-2 (SGLT2) inhibitors,^26–28^ mineralocorticoid receptor antagonists,^29–31^ glucagon-like peptide-1 receptor agonists,^32^ and ACEi/ARB,^33^ is critical in those with evidence of cardiovascular or kidney disease.

Cardiovascular, kidney, and metabolic disorders arise from complex interactions between shared risk factors.^2^ There is likely heterogeneity within each CKM stage, leading some individuals to progress, whereas others do not. Furthermore, depending on the interplay of risk factors, a patient may progress towards cardiovascular disease, kidney disease, or both. Identifying distinct phenotypes within each stage will be an important step towards explaining this variability. Further work is needed to elucidate the interactions between cardiovascular risk factors and develop strategies to mitigate them. The AHA’s Presidential Advisory on CKM syndrome challenges us to consider a spectrum of disease processes that are inextricably linked to environmental, genetic, and social risks under a unifying conceptual framework of cardiovascular risk.^1^ Ultimately, clinical co-management in the form of multi-disciplinary clinics may improve outcomes for those already affected by CKM syndrome.^34^ However, identifying and staging individuals within CKM syndrome according to the AHA advisory will require increased vigilance by frontline primary care and community health workers.

Strengths of this study include the use of a nationally representative sample, long duration of follow-up, comprehensive covariate assessment, and low rates of missing data. The analyses would be strengthened by inclusion of incident major cardiovascular events; however, these data are not available in NHANES. Specific limitations related to defining CKM syndrome stages in NHANES are described in **Table S15**.

In conclusion, CKM syndrome stages were associated with a graded risk of cardiovascular mortality in this nationally representative sample of U.S. adults. Stages 1 and 2 of CKM syndrome are highly prevalent in the U.S. population, and this emphasizes the need for increased obesity treatment and focused preventative efforts. Our findings provide a foundational view of CKM syndrome in the U.S. and offer guideposts for improving the treatment of intertwined metabolic, kidney, and cardiovascular diseases in the U.S.

## Data Availability

All data are available at https://wwwn.cdc.gov/nchs/nhanes/default.aspx

