## Supplementary material for "Prevalence and Cumulative Incidence of Mortality Associated with Cardiovascular-Kidney-Metabolic Syndrome in the United States": Table S1

**Supplemental Table 1. Variable Definitions**

| **Variable** | **Definition** |
|---|---|
| Hypertension | Systolic blood pressure ≥130 mmHg OR diastolic blood pressure ≥80 mmHg OR current use of an antihypertensive medication |
| Diabetes | Hemoglobin A1c (HbA1c) ≥6.5% (44.0 mmol/mol) OR current use of insulin OR current use of an anti-hyperglycemic medication |
| Obesity | Body mass index (BMI) ≥30 kg/m$^2$ |
| Hyperlipidemia | Total cholesterol ≥240 mg/dL (6.5 mmol/L) OR current use of a lipid-lowering medication |
| Chronic Kidney Disease* | |
| None | [eGFR ≥90 ml/min per 1.73m$^2$ and UACR < 30 mg/g (< 3 mg/mmol)] OR [eGFR 60-90 ml/min per 1.73m$^2$ and UACR < 30 mg/g (< 3 mg/mmol)] |
| Low risk | [eGFR ≥90 ml/min per 1.73m$^2$ and UACR 30-300 mg/g (3-30 mg/mmol)] OR [eGFR 60-90 ml/min per 1.73m$^2$ and UACR 30-300 mg/g (3-30 mg/mmol)] OR [eGFR 45-60 ml/min per 1.73m$^2$ and UACR < 30 mg/g (< 3 mg/mmol)] |
| Moderate to high risk | [eGFR ≥90 ml/min per 1.73m$^2$ and UACR ≥300 mg/g (≥30 mg/mmol)] OR [eGFR 60-90 ml/min per 1.73m$^2$ and UACR ≥300 mg/g (≥30 mg/mmol)] OR [eGFR 45-60 ml/min per 1.73m$^2$ and UACR 30-300 mg/g (3-30 mg/mmol)] OR [eGFR 30-45 ml/min per 1.73m$^2$ and UACR < 30 mg/g (< 3 mg/mmol)] |
| Very high risk | [eGFR 45-60 ml/min per 1.73m$^2$ and UACR ≥300 mg/g (≥30 mg/mmol)] OR [eGFR 30-45 ml/min per 1.73m$^2$ and UACR 30-300 mg/g (3-30 mg/mmol)] OR [eGFR 30-45 ml/min per 1.73m$^2$ and UACR ≥300 mg/g (≥30 mg/mmol)] OR [eGFR ≤ 30 ml/min per 1.73m$^2$] |
| Metabolic Syndrome | Presence of at least 3 of the following: (1) waist circumference ≥88/102 cm in women/men; (2) HDL <40 mg/dL for men and <50 mg/dL for women; (3) hypertension; (4) HbA1c ≥5.7% (38.8 mmol/mol) |
| History of cardiovascular disease | Ever been told by a medical professional that you have had coronary heart disease, angina pectoris, heart failure, a stroke, or a heart attack. |
| History of cancer | Ever been told by a medical professional that you have had a cancer or malignancy of any kind. |
| History of respiratory disease | Ever been told by a medical profession that you have chronic bronchitis, emphysema, asthma, or chronic obstructive pulmonary disease. |
| History of liver disease | Ever been told by a medical professional that you have had any kind of liver condition. |
| CKM Stages[†] | |
| Stage 0 | BMI <25 kg/m$^2$ AND HbA1c <5.7% (38.8 mmol/mol) AND absent insulin use, anti-hyperglycemic medication use, hypertension, chronic kidney disease, hyperlipidemia, and cardiovascular disease. |
| Stage 1 | (BMI ≥25 kg/m$^2$ OR HbA1c 5.7-6.4% (38.8-46.4 mmol/mol) OR waist circumference ≥88/102 cm in women/men[‡]) AND absent hypertension, insulin use, anti-hyperglycemic medication use, low-risk chronic kidney disease, hyperlipidemia, and cardiovascular disease. |
| Stage 2 | (Metabolic syndrome OR hypertension OR diabetes OR moderate- to high-risk chronic kidney disease OR hyperlipidemia) AND absent cardiovascular disease. |
| Stage 3 | Very high-risk chronic kidney disease OR 10-year PREVENT risk score ≥20% AND absent cardiovascular disease. |
| Stage 4 | Prevalent cardiovascular disease. |

* Based on the Kidney Disease Improving Global Outcomes (KDIGO) 2022 chronic kidney disease heatmap.

[†] CKM Stage Definitions from Ndumele *et al.* 2023:

Stage 0: includes individuals with normal weight, normal glucose, normal blood pressure, normal lipids, normal kidney function, and no evidence of subclinical or clinical cardiovascular disease.

Stage 1: includes individuals with excess adipose tissue, dysfunctional adipose tissue, or both. Excess adiposity is identified by either weight or abdominal obesity, and dysfunctional adipose tissue is reflected by impaired glucose tolerance and hyperglycemia.

Stage 2: includes individuals with metabolic risk factors (hypertriglyceridemia, hypertension, metabolic syndrome, or type 2 diabetes), moderate- to high-risk chronic kidney disease, or both.

Stage 3: includes individuals with subclinical cardiovascular disease with overlapping CKM risk factors (excess/dysfunctional adipose tissue, metabolic risk factors, or chronic kidney disease) or those with the risk equivalents of very high-risk chronic kidney disease or high 10-year predicted cardiovascular risk.

Stage 4: includes individuals with clinical cardiovascular disease (coronary heart disease, heart failure, stroke, peripheral artery disease, or atrial fibrillation) overlapping with CKM risk factors.

| **Supplemental Table 2. Missingness** | | |
|---|---|---|
| | **1999-2018 Sample N (%)** | **1999-2020 Sample N (%)** |
| Systolic blood pressure | 1,973 (3.9) | 2,103 (3.9) |
| Urinary albumin: creatinine ratio | 1,320 (2.6) | 1,397 (2.6) |
| Waist circumference | 993 (2.0) | 1,606 (3.0) |
| Body mass index | 808 (1.6) | 851 (1.1) |
| Serum creatinine | 154 (0.3) | 3,551 (6.6) |
| Hemoglobin A1c | 69 (0.1) | 72 (0.1) |
| Total cholesterol | 3,087 (6.1) | 3,398 (6.3) |
| Food security | 1,005 (2.0) | |
| Education | 43 (0.08) | |
| Poverty-to-Income ratio | 3,963 (7.8) | |
| Health insurance | 171 (0.3) | |
| History of cancer | 41 (0.08) | |
| History of liver disease | 86 (0.2) | |

| Supplemental Table 3. Participant Characteristics Prior to Imputation | | | | | | |
|---|---|---|---|---|---|---|
| | **Overall** | **Stage 0** | **Stage 1** | **Stage 2** | **Stage 3** | **Stage 4** |
| No. participants * | 46,058 | 4,995 | 6,989 | 18,661 | 10,405 | 5,008 |
| Age, years | 47.1 (16.7) | 35.4 (12.3) | 39.8 (13.0) | 55.1 (14.7) | 40.2 (14.1) | 64.4 (13.6) |
| Female sex | 51.1 | 64.1 | 66.2 | 66.3 | 12.4 | 45.7 |
| Race or Ethnicity † | | | | | | |
| Non-Hispanic White | 68.9 | 70.9 | 63.6 | 70.5 | 66.6 | 75.8 |
| Non-Hispanic Black | 8.2 | 6.7 | 11.3 | 6.2 | 11.3 | 4.2 |
| Mexican American | 5.6 | 5.5 | 7.2 | 4.8 | 6.6 | 3.4 |
| Other Hispanic | 10.5 | 7.7 | 12.5 | 12.2 | 8.1 | 10.7 |
| Other Race | 6.8 | 9.2 | 5.5 | 6.3 | 7.4 | 5.9 |
| Poverty-to-Income Ratio | | | | | | |
| <100% FPL | 13.6 | 13.5 | 15.1 | 11.9 | 14.5 | 16.5 |
| 100-300% FPL | 36.1 | 32.3 | 34.6 | 36.1 | 36.0 | 45.8 |
| ≥300% FPL | 50.2 | 54.2 | 50.3 | 52.1 | 49.5 | 37.7 |
| Education | | | | | | |
| Below High School | 17.1 | 11.5 | 15.1 | 17.3 | 17.8 | 26.3 |
| High School/GED | 23.9 | 19.2 | 20.9 | 25.0 | 25.8 | 27.2 |
| Above High School | 59.0 | 69.3 | 64.0 | 57.8 | 56.4 | 46.5 |
| Uninsured | 17.7 | 21.3 | 21.2 | 12.8 | 23.9 | 8.7 |
| Food insecurity | 13.4 | 12.0 | 14.6 | 11.7 | 14.9 | 16.1 |
| PREVENT Score Categories | | | | | | |
| Low Risk (<5%) | 70.1 | 96.6 | 94.2 | 95.6 | 1.7 | 67.2 |
| Borderline Risk (5-7.4%) | 0.9 | 1.1 | 1.8 | 1.1 | 0 | 0.7 |
| Intermediate Risk (7.5-19.9%) | 2.4 | 2.2 | 4.1 | 3.3 | 0.04 | 2.3 |
| High Risk (≥20%) | 26.5 | 0 | 0 | 0 | 98.3 | 29.8 |
| Current smoking | 21.5 | 23.6 | 19.3 | 17.9 | 27.4 | 22.1 |
| Diabetes | 10.3 | 0 | 0 | 13.4 | 11.4 | 29.9 |
| Hypertension | 48.1 | 0 | 0 | 78.1 | 47.2 | 85.5 |
| Hyperlipidemia | 31.5 | 0 | 0 | 41.1 | 41.9 | 69.5 |
| Obesity | 35.4 | 0 | 35.9 | 39.2 | 44.8 | 45.3 |
| Metabolic syndrome | 24.7 | 0 | 0 | 35.8 | 28.4 | 51.6 |
| Medical History | | | | | | |
| Chronic respiratory disease | 15.3 | 14.3 | 15.8 | 16.2 | 14.2 | 28.4 |
| Liver disease | 3.5 | 1.7 | 2.1 | 4.1 | 3.2 | 7.2 |
| Cancer | 9.3 | 4.8 | 5.1 | 12.6 | 5.3 | 21.3 |
| Chronic kidney disease | 13.1 | 0 | 0 | 19.9 | 10.1 | 37.0 |

| | | | | | | |
|---|---|---|---|---|---|---|
| Cardiovascular disease | 8.4 | 0 | 0 | 0 | 0 | 99.7 |
| Cardioprotective Medication Use | | | | | | |
| Statin | 12.8 | 0 | 0 | 22.1 | 12.6 | 40.1 |
| ACEi/ARB | 14.8 | 0 | 0 | 12.0 | 19.7 | 40.9 |
| Hemoglobin A1c, % | 5.3 [5.1, 5.7] | 5.1 [4.9, 5.3] | 5.2 [5.0, 5.5] | 5.5 [5.2, 5.8] | 5.3 [5.1, 5.7] | 5.7 [5.3, 6.2] |
| Total Cholesterol, mg/dL | 196.3 (41.7) | 175.1 (29.4) | 182.1 (29.4) | 204.0 (42.0) | 208.8 (44.6) | 186.7 (46.0) |
| High Density Lipoprotein, mg/dL | 53.1 (16.2) | 62.1 (14.6) | 55.6 (13.2) | 57.3 (17.0) | 41.1 (9.3) | 50.2 (16.2) |
| BMI, $kg/m^2$ | 28.8 (6.7) | 21.8 (2.0) | 29.7 (5.4) | 29.5 (7.1) | 30.2 (6.2) | 30.3 (7.0) |
| Waist circumference, cm | | | | | | |
| Women | 95.5 (11.6) | 77.4 (4.4) | 97.4 (10.0) | 98.8 (13.1) | 104.5 (5.8) | 102.3 (10.8) |
| Men | 101.2 (10.7) | 82.7 (4.1) | 99.6 (6.7) | 102.2 (8.8) | 103.7 (13.8) | 107.6 (10.7) |
| Systolic blood pressure, mmHg | 122.7 (17.4) | 109.3 (9.1) | 112.3 (8.7) | 130.0 (18.4) | 122.5 (14.4) | 130.7 (20.9) |
| eGFR, ml/min per $1.73m^2$ | 96.1 (20.4) | 106.4 (16.1) | 103.1 (16.5) | 90.5 (18.8) | 100.9 (19.5) | 77.6 (23.1) |
| UACR, mg/g | 6.4 [4.2, 11.6] | 5.7 [4.1, 8.8] | 5.3 [3.8, 8.1] | 4.9 [7.7, 15.5] | 5.4 [3.7, 9.6] | 10.0 [5.9, 28.6] |

* Sample size for each CKM stage is unweighted, but all other numbers in the table are weighted percentages, means (with standard deviations), or medians (with interquartile range).
† Race or ethnicity was self-reported by participants.

Abbreviations: ACEi, angiotensin converting enzyme inhibitor; ARB, angiotensin receptor blocker; BMI, body mass index; eGFR, estimated glomerular filtration rate; FPL, federal poverty level; GED, general education diploma; PREVENT, predicting risk of cardiovascular disease events; UACR, urinary albumin-to-creatinine ratio.

| Supplemental Table 4. Trends in Age-Adjusted* Prevalence of CKM Syndrome, 1999-2020. | | | | | | | |
|---|---|---|---|---|---|---|---|
| | **Overall** | **1999-2002** | **2003-2006** | **2007-2010** | **2011-2014** | **2015-2020** | ***p*-trend** |
| Stage 0 | 12.5 (12.0, 12.9) | 13.5 (12.2, 14.8) | 13.8 (12.8, 14.8) | 12.2 (11.5, 13.0) | 11.8 (10.8, 12.8) | 11.4 (10.2, 12.6) | 0.002 |
| Stage 1 | 16.7 (16.2, 17.2) | 13.6 (12.3, 14.9) | 15.4 (14.1, 16.6) | 16.6 (15.3, 17.9) | 17.8 (16.7, 19.0) | 19.1 (18.1, 20.0) | <0.001 |
| Stage 2 | 40.0 (38.4, 39.6) | 38.9 (37.7, 40.0) | 38.5 (37.3, 39.8) | 38.0 (36.7, 39.4) | 39.1 (37.9, 40.4) | 40.0 (38.7, 41.4) | 0.11 |
| Stage 3 | 22.9 (22.5, 23.4) | 24.8 (23.7, 25.9) | 22.4 (21.5, 23.4) | 24.7 (23.6, 25.8) | 22.7 (21.5, 23.9) | 20.7 (19.6, 21.7) | <0.001 |
| Stage 4 | 8.9 (8.6, 9.2) | 9.2 (8.3, 10.1) | 9.8 (8.9, 10.8) | 8.5 (7.7, 9.2) | 8.5 (7.8, 9.1) | 8.8 (8.2, 9.5) | 0.11 |
| * Age-standardized to the 2010 U.S. Census | | | | | | | |

| Supplemental Table 5. Trends in Age-Adjusted* Prevalence of CKM Syndrome by Sex, 1999-2020. | | | | | | |
|---|---|---|---|---|---|---|
| | **1999-2002** | **2003-2006** | **2007-2010** | **2011-2014** | **2015-2020** | ***p*-trend** |
| **Women** | | | | | | |
| Stage 0 | 17.8 (16.0, 19.6) | 18.1 (16.4, 19.7) | 16.2 (14.9, 17.4) | 14.3 (13.0, 15.6) | 13.5 (12.0, 15.1) | <0.001 |
| Stage 1 | 19.6 (17.8, 21.4) | 19.6 (17.9, 21.3) | 21.4 (19.9, 22.9) | 22.7 (21.4, 24.0) | 24.2 (22.5, 25.9) | <0.001 |
| Stage 2 | 47.9 (46.1, 49.7) | 48.1 (46.6, 49.6) | 48.6 (46.8, 50.5) | 49.6 (47.8, 51.5) | 50.2 (48.7, 51.6) | 0.02 |
| Stage 3 | 6.9 (6.0, 7.7) | 5.2 (4.3, 6.1) | 6.8 (5.8, 7.8) | 5.8 (4.8, 6.8) | 4.4 (3.8, 5.0) | <0.001 |
| Stage 4 | 7.8 (6.8, 8.9) | 9.1 (7.9, 10.3) | 7.0 (6.1, 8.0) | 7.6 (6.6, 8.5) | 7.7 (6.8, 8.5) | 0.24 |
| **Men** | | | | | | |
| Stage 0 | 9.4 (8.1, 10.8) | 9.7 (8.5, 11.0) | 8.4 (7.3, 9.5) | 9.3 (8.2, 10.5) | 9.3 (7.8, 10.7) | 0.84 |
| Stage 1 | 7.7 (6.5, 8.9) | 11.4 (10.0, 12.7) | 11.9 (10.4, 13.3) | 13.0 (11.7, 14.3) | 14.0 (12.8, 15.3) | <0.001 |
| Stage 2 | 29.2 (27.6, 30.8) | 28.2 (26.8, 29.7) | 26.5 (25.0, 27.9) | 27.8 (26.1, 29.4) | 29.1 (27.3, 30.8) | 0.78 |
| Stage 3 | 42.9 (40.9, 44.8) | 39.8 (38.1, 41.5) | 43.1 (41.4, 44.7) | 40.3 (38.3, 42.3) | 37.4 (35.8, 39.1) | <0.001 |
| Stage 4 | 10.9 (9.6, 12.1) | 10.9 (9.7, 12.0) | 10.2 (9.3, 11.1) | 9.6 (8.7, 10.5) | 10.2 (9.4, 11.1) | 0.17 |
| * Age-standardized to the 2010 U.S. Census | | | | | | |

| Supplemental Table 6. Trends in Prevalence of CKM Syndrome by Age Group, 1999-2020. | | | | | | |
|---|---|---|---|---|---|---|
| | **1999-2002** | **2003-2006** | **2007-2010** | **2011-2014** | **2015-2020** | ***p*-trend** |
| **Age 20-39 years** | | | | | | |
| Stage 0 | 22.6 (20.2, 25.0) | 26.0 (24.0, 28.1) | 22.4 (20.7, 24.0) | 22.0 (20.0, 24.0) | 21.5 (19.1, 24.0) | 0.11 |
| Stage 1 | 19.1 (17.2, 20.9) | 21.8 (19.7, 23.9) | 23.6 (21.0, 26.2) | 26.7 (24.5, 28.9) | 27.6 (25.8, 29.4) | <0.001 |
| Stage 2 | 17.8 (16.2, 19.4) | 18.5 (17.2, 19.8) | 16.5 (15.0, 18.1) | 18.0 (15.9, 20.2) | 20.8 (19.1, 22.4) | 0.02 |
| Stage 3 | 39.4 (37.3, 41.5) | 32.7 (30.5, 34.9) | 36.4 (34.2, 38.5) | 31.8 (29.3, 34.4) | 28.8 (26.8, 30.8) | <0.001 |
| Stage 4 | 1.2 (0.8, 1.5) | 1.00 (0.6, 1.4) | 1.1 (0.7, 1.5) | 1.4 (1.0, 1.9) | 1.3 (0.9, 1.7) | 0.29 |
| **Age 40-59 years** | | | | | | |
| Stage 0 | 11.8 (9.9, 13.7) | 9.9 (8.3, 11.5) | 9.4 (8.0, 10.8) | 8.3 (7.0, 9.5) | 7.6 (6.3, 8.9) | <0.001 |
| Stage 1 | 14.4 (12.5, 16.2) | 15.7 (13.8, 17.7) | 17.2 (15.5, 18.9) | 17.8 (16.3, 19.3) | 19.3 (17.6, 21.1) | <0.001 |
| Stage 2 | 44.1 (41.7, 46.4) | 45.3 (42.7, 47.8) | 44.7 (42.4, 47.0) | 45.9 (43.3, 48.5) | 46.6 (44.3, 48.8) | 0.13 |
| Stage 3 | 23.0 (21.1, 24.9) | 22.2 (20.5, 24.0) | 22.9 (21.4, 24.5) | 22.4 (20.3, 24.5) | 19.7 (18.1, 21.3) | 0.01 |
| Stage 4 | 6.8 (5.3, 8.3) | 6.8 (5.4, 8.2) | 5.7 (5.0, 6.5) | 5.7 (4.6, 6.8) | 6.8 (5.7, 7.9) | 0.70 |
| **Age ≥60 years** | | | | | | |
| Stage 0 | 3.0 (2.3, 3.8) | 2.0 (1.5, 2.5) | 1.8 (1.1, 25.5) | 2.3 (1.6, 3.0) | 2.3 (1.5, 3.2) | 0.59 |
| Stage 1 | 4.3 (3.4, 5.2) | 5.5 (4.3, 6.7) | 5.3 (4.4, 6.3) | 5.1 (3.9, 6.3) | 6.3 (5.2, 7.4) | <0.001 |
| Stage 2 | 61.8 (59.4, 64.1) | 57.5 (55.6, 59.3) | 59.3 (57.5, 61.0) | 59.6 (57.4, 61.7) | 58.2 (56.1, 60.3) | 0.18 |
| Stage 3 | 6.5 (5.7, 7.2) | 7.9 (6.7, 9.1) | 10.5 (9.3, 11.7) | 10.1 (8.8, 11.4) | 10.4 (9.0, 11.7) | <0.001 |
| Stage 4 | 24.5 (22.1, 26.8) | 27.1 (25.0, 29.3) | 23.1 (20.8, 25.5) | 22.9 (21.0, 24.8) | 22.8 (21.1, 24.6) | 0.02 |

| Supplemental Table 7. Age-Adjusted* Prevalence of CKM Syndrome by Race or Ethnicity. | | | | | |
|---|---|---|---|---|---|
| | **Stage 0** | **Stage 1** | **Stage 2** | **Stage 3** | **Stage 4** |
| **Non-Hispanic White** | 13.9 (13.2, 14.6) | 16.0 (15.3, 16.7) | 37.7 (37.0, 38.5) | 23.4 (22.8, 24.1) | 8.9 (8.5, 9.4) |
| **Hispanic‡** | 9.2 (8.6, 9.9) | 19.1 (18.3, 19.9) | 38.6 (37.7, 39.5) | 25.9 (24.9, 26.9) | 7.1 (6.6, 7.6) |
| **Non-Hispanic Black** | 8.2 (7.7, 8.7) | 17.6 (16.8, 18.4) | 48.1 (47.1, 49.0) | 15.9 (15.2, 16.6) | 10.3 (9.7, 10.9) |
| **Other Race** | 12.8 (11.1, 14.4) | 13.9 (11.9, 16.0) | 38.0 (35.5, 40.5) | 24.1 (22.0, 26.2) | 11.3 (9.6, 13.0) |
| **Non-Hispanic Asian†** | 15.3 (13.6, 16.9) | 17.1 (15.7, 18.6) | 39.9 (38.1, 41.7) | 22.4 (20.9, 24.0) | 5.3 (4.3, 6.3) |

* Age-standardized to the 2010 U.S. Census.

‡ Hispanic includes both the ‘Mexican American’ and ‘Other Hispanic’ categories.

† Non-Hispanic Asian was first reported as a unique race category in the 2011-2012 survey year. Therefore, estimates are only representative of 2011-2020 for Non-Hispanic Asians. Prior to 2011, Non-Hispanic Asians were included in ‘Other Race.’

| Supplemental Table 8. Trends in Age-Adjusted* Prevalence of CKM syndrome by Race or Ethnicity, 1999-2020. | | | | | | |
|---|---|---|---|---|---|---|
| | **1999-2002** | **2003-2006** | **2007-2010** | **2011-2014** | **2015-2020** | ***p*-trend** |
| **Non-Hispanic White** | | | | | | |
| Stage 0 | 14.9 (13.2, 16.6) | 15.1 (13.7, 16.6) | 13.5 (12.3, 14.7) | 13.0 (11.5, 14.4) | 13.3 (11.5, 15.1) | 0.06 |
| Stage 1 | 12.9 (11.2, 14.5) | 14.8 (13.1, 16.4) | 16.1 (14.4, 17.8) | 17.2 (15.4, 18.9) | 18.5 (17.1, 19.9) | <0.001 |
| Stage 2 | 37.1 (35.5, 38.8) | 37.3 (35.8, 38.8) | 37.3 (35.5, 39.0) | 38.1 (36.4, 39.8) | 38.8 (36.9, 40.6) | 0.22 |
| Stage 3 | 25.8 (24.6, 26.9) | 22.9 (21.6, 24.3) | 24.9 (23.5, 26.3) | 23.4 (21.8, 25.0) | 20.5 (19.1, 22.0) | <0.001 |
| Stage 4 | 9.4 (8.4, 10.4) | 9.9 (8.9, 10.9) | 8.3 (7.3, 9.3) | 8.4 (7.5, 9.3) | 8.9 (8.0, 9.8) | 0.12 |
| **Hispanic‡** | | | | | | |
| Stage 0 | 11.0 (8.8, 13.1) | 11.0 (9.4, 12.6) | 8.9 (7.8, 10.0) | 8.7 (7.3, 10.1) | 7.9 (7.0, 8.8) | 0.001 |
| Stage 1 | 17.6 (15.7, 19.6) | 17.9 (15.9, 19.8) | 18.2 916.5, 19.9) | 20.5 (18.9, 22.1) | 20.4 (18.8, 21.9) | <0.001 |
| Stage 2 | 38.8 (35.9, 41.6) | 38.8 (36.8, 40.9) | 36.7 (35.3, 38.2) | 38.2 (36.9, 39.4) | 40.1 (39.2, 42.1) | 0.10 |
| Stage 3 | 25.6 (23.2, 28.1) | 25.5 (23.2, 27.7) | 29.1 (26.9, 31.4) | 25.2 (23.4, 26.9) | 24.6 (22.8, 26.5) | 0.05 |
| Stage 4 | 7.0 (5.8, 8.2) | 6.9 (5.8, 7.9) | 7.0 (6.0, 8.0) | 7.4 (6.3, 8.6) | 7.0 (6.0, 7.9) | 0.97 |
| **Non-Hispanic Black** | | | | | | |
| Stage 0 | 9.3 (7.8, 10.8) | 8.2 (6.8, 9.5) | 8.5 (7.4, 9.5) | 7.7 (6.9, 8.6) | 7.6 (6.7, 8.6) | 0.09 |
| Stage 1 | 14.6 (13.2, 16.0) | 17.4 (15.1, 19.7) | 18.5 (16.7, 20.2) | 18.0 (16.7, 19.3) | 18.7 (17.1, 20.4) | 0.001 |
| Stage 2 | 48.0 (46.0, 50.0) | 46.9 (44.3, 49.4) | 47.0 (44.9, 49.1) | 47.5 (45.6, 49.4) | 50.0 (48.0, 51.9) | 0.12 |
| Stage 3 | 17.8 (15.8, 19.7) | 16.7 (15.7, 17.7) | 15.8 (14.2, 17.3) | 16.9 (15.4, 18.4) | 13.4 (12.1, 14.7) | <0.001 |
| Stage 4 | 10.4 (8.9, 11.9) | 10.9 (9.4, 12.3) | 10.3 (9.0, 11.6) | 10.0 (8.9, 11.0) | 10.3 (9.2, 11.4) | 0.80 |
| **Other Race** | | | | | | |
| Stage 0 | 10.8 (5.8, 15.8) | 18.1 (13.7, 22.6) | 14.9 (12.2, 17.6) | 15.3 (13.2, 17.3) | 11.2 (9.5, 13.0) | 0.18 |
| Stage 1 | 7.9 (3.9, 11.9) | 11.2 (8.0, 14.4) | 13.7 (9.5, 17.9) | 16.3 (14.0, 18.5) | 18.8 (16.6, 21.1) | <0.001 |
| Stage 2 | 47.3 (40.5, 54.2) | 39.3 (33.4, 45.2) | 35.0 (29.9, 10.1) | 38.8 (36.6, 41.1) | 37.7 (35.5, 40.0) | 0.04 |
| Stage 3 | 26.3 (19.4, 33.3) | 20.3 (16.0, 24.5) | 27.8 (24.2, 31.5) | 21.5 (19.1, 24.0) | 23.1 (20.9, 25.2) | 0.36 |
| Stage 4 | 7.6 (3.7, 11.5) | 11.1 (6.3, 15.8) | 8.5 (6.3, 10.8) | 8.1 (6.3, 9.9) | 9.2 (7.3, 11.0) | 0.94 |

* Age-standardized to the 2010 U.S. Census
‡ Hispanic includes both the ‘Mexican American’ and ‘Other Hispanic’ categories.

| **Supplemental Table 9. Trends in Age-Adjusted* Prevalence of CKM syndrome by Race or Ethnicity, 2011-2020.** | | | | | |
|---|---|---|---|---|---|
| | **2011-2012** | **2013-2014** | **2015-2016** | **2017-2020** | ***p*-trend** |
| **Non-Hispanic White** | | | | | |
| Stage 0 | 12.3 (9.9, 14.6) | 13.7 (12.1, 15.4) | 13.2 (10.9, 15.5) | 13.3 (10.8, 15.9) | 0.65 |
| Stage 1 | 16.2 (14.3, 18.0) | 18.2 (15.3, 21.1) | 19.4 (17.2, 21.6) | 18.0 (16.1, 19.8) | 0.18 |
| Stage 2 | 40.4 (37.6, 43.2) | 35.8 (34.3, 37.3) | 37.4 (35.0, 69.9) | 39.6 (36.9, 42.2) | 0.97 |
| Stage 3 | 22.9 (21.4, 24.5) | 23.8 (20.9, 26.6) | 22.3 (20.1, 24.5) | 19.5 (17.4, 21.5) | 0.01 |
| Stage 4 | 8.2 (7.3, 9.1) | 8.6 (7.1, 10.1) | 7.7 (6.7, 8.7) | 9.7 (8.4, 11.0) | 0.14 |
| **Hispanic[‡]** | | | | | |
| Stage 0 | 8.5 (6.8, 10.2) | 9.0 (6.7, 11.2) | 6.8 (5.5, 8.2) | 8.5 (7.4, 9.7) | 0.70 |
| Stage 1 | 20.0 (17.3, 22.7) | 21.0 (19.2, 22.8) | 20.1 (17.8, 22.4) | 20.5 (18.5, 22.5) | 0.94 |
| Stage 2 | 37.3 (35.3, 39.3) | 38.9 (37.8, 40.1) | 39.5 (37.1, 41.9) | 40.5 (37.8, 43.3) | 0.02 |
| Stage 3 | 25.9 (22.9, 28.8) | 24.5 (22.5, 26.5) | 26.4 (23.8, 29.0) | 23.6 (21.1, 26.0) | 0.23 |
| Stage 4 | 8.4 (6.6, 10.2) | 6.6 (5.5, 7.7) | 7.2 (5.9, 8.5) | 6.9 (5.6, 8.2) | 0.23 |
| **Non-Hispanic Black** | | | | | |
| Stage 0 | 7.2 (6.2, 8.3) | 8.2 (6.9, 9.6) | 8.1 (6.2, 9.9) | 7.4 (6.3, 8.5) | 0.94 |
| Stage 1 | 17.8 (16.0, 19.6) | 18.2 (16.3, 20.1) | 19.8 (16.7, 23.0) | 18.1 (16.1, 20.0) | 0.76 |
| Stage 2 | 46.8 (43.7, 49.9) | 48.2 (45.8, 50.5) | 47.4 (44.2, 50.7) | 51.5 (49.1, 54.0) | 0.02 |
| Stage 3 | 17.8 (15.8, 19.7) | 16.0 (13.7, 18.3) | 14.0 (12.4, 15.5) | 13.0 (11.1, 14.8) | <0.001 |
| Stage 4 | 10.5 (8.7, 12.2) | 9.4 (8.3, 10.6) | 10.7 (9.1, 12.4) | 10.1 (8.6, 11.5) | 0.98 |
| **Non-Hispanic Asian[†]** | | | | | |
| Stage 0 | 17.3 (14.2, 20.3) | 18.4 (14.7, 22.1) | 15.5 (12.5, 18.5) | 12.3 (9.9, 14.7) | 0.003 |
| Stage 1 | 16.3 (13.3, 19.4) | 17.1 (13.2, 21.0) | 18.2 (16.4, 20.0) | 16.9 (14.5, 19.4) | 0.77 |
| Stage 2 | 41.2 (38.1, 44.4) | 37.0 (33.3, 40.6) | 39.8 (37.2, 42.3) | 40.9 (37.4, 44.4) | 0.55 |
| Stage 3 | 20.4 (17.4, 23.4) | 21.6 (17.6, 25.6) | 22.0 (19.5, 24.5) | 24.2 (21.4, 27.0) | 0.06 |
| Stage 4 | 4.8 (3.6, 6.0) | 6.0 (4.6, 7.3) | 4.5 (2.5, 6.6) | 5.7 (3.7, 7.6) | 0.65 |
| **Other Race** | | | | | |
| Stage 0 | 9.2 (6.2, 12.3) | 12.0 (7.0, 17.1) | 9.2 (1.9, 16.4) | 6.9 (4.2, 9.6) | 0.14 |
| Stage 1 | 16.2 (9.6, 22.7) | 14.2 (4.8, 23.6) | 22.6 (13.6, 31.6) | 20.3 (14.3, 26.3) | 0.19 |
| Stage 2 | 42.5 (36.0, 49.0) | 31.5 (20.3, 42.7) | 28.9 (22.8, 35.0) | 36.1 (30.3, 41.9) | 0.25 |

| | | | | | |
|---|---|---|---|---|---|
| Stage 3 | 16.4 (9.9, 22.9) | 29.4 (21.6, 37.3) | 20.7 (13.2, 28.2) | 24.1 (18.4, 29.9) | 0.29 |
| Stage 4 | 15.7 (10.5, 21.0) | 12.9 (4.7, 21.1) | 18.7 (12.1, 25.3) | 12.6 (8.8, 16.3) | 0.50 |

* Age-standardized to the 2010 U.S. Census

‡ Hispanic includes both the 'Mexican American' and 'Other Hispanic' categories.

† Non-Hispanic Asian was first reported as a unique race category in the 2011-2012 survey year.

| Supplemental Table 10. Association between CKM syndrome stage and mortality, adjusting for Social Determinants of Health | | | | | |
|---|---|---|---|---|---|
| **CVD Mortality** | **Stage 0** | **Stage 1** | **Stage 2** | **Stage 3** | **Stage 4** |
| No. events | 27 | 60 | 1,143 | 256 | 1,071 |
| No. participants | 5,174 | 7,544 | 20,838 | 11,243 | 5,825 |
| Multivariable model 1[a] | Referent | 1.06 [0.63, 1.79] | 1.60 [0.96, 2.67] | 1.90 [1.10, 3.25] | 3.93 [2.30, 6.70] |
| Multivariable model 2[b] | Referent | 1.03 [0.61, 1.74] | 1.56 [0.93, 2.61] | 1.85 [1.08, 3.18] | 3.80 [2.22, 6.50] |
| Multivariable model 3[c] | Referent | 1.03 [0.61, 1.73] | 1.55 [0.92, 2.59] | 1.83 [1.07, 3.16] | 3.72 [2.17, 6.37] |
| Multivariable model 4[d] | Referent | 1.02 [0.61, 1.73] | 1.52 [0.91, 2.55] | 1.79 [1.05, 3.07] | 3.61 [2.11, 6.18] |
| Multivariable model 5[e] | Referent | 1.04 [0.62, 1.74] | 1.52 [0.91, 2.55] | 1.77 [1.03, 3.04] | 3.51 [2.04, 6.02] |
| **All-Cause Mortality** | **Stage 0** | **Stage 1** | **Stage 2** | **Stage 3** | **Stage 4** |
| No. events | 207 | 347 | 3,942 | 1,027 | 2,646 |
| No. participants | 5,174 | 7,544 | 20,838 | 11,243 | 5,825 |
| Multivariable model 1[a] | Referent | 0.98 [0.81, 1.19] | 1.14 [0.96, 1.36] | 1.46 [1.20, 1.77] | 2.06 [1.72, 2.48] |
| Multivariable model 2[b] | Referent | 0.97 [0.80, 1.19] | 1.14 [0.95, 1.37] | 1.44 [1.17, 1.77] | 2.05 [1.69, 2.49] |
| Multivariable model 3[c] | Referent | 0.96 [0.79, 1.18] | 1.13 [0.94, 1.35] | 1.42 [1.16, 1.75] | 2.00 [1.65, 2.42] |
| Multivariable model 4[d] | Referent | 0.96 [0.78, 1.18] | 1.11 [0.92, 1.33] | 1.38 [1.12, 1.70] | 1.93 [1.59, 2.35] |
| Multivariable model 5[e] | Referent | 0.97 [0.79, 1.18] | 1.10 [0.92, 1.32] | 1.35 [1.10, 1.67] | 1.87 [1.53, 2.27] |

[a] Multivariable model 1: adjusted for age, sex, race or ethnicity, smoking status, survey year, BMI, waist circumference, hemoglobin A1c, systolic blood pressure, total cholesterol, statin use, ACE/ARB use, UACR, eGFR, history of chronic respiratory disease, history of liver disease, history of cancer.
[b] Multivariable model 2: adjusted for Model 1 + health insurance.
[c] Multivariable model 3: adjusted for Model 2 + food security.
[d] Multivariable model 4: adjusted for Model 3 + educational attainment.
[e] Multivariable model 5: adjusted for Model 4 + poverty-to-income ratio.

| **Supplemental Table 11. Association between CKM syndrome stage and cardiovascular mortality** | | | | | |
|---|---|---|---|---|---|
| **Unweighted Models to Contextualize the Unweighted Adjusted Cumulative Incidence Estimates** | | | | | |
| | **Stage 0** | **Stage 1** | **Stage 2** | **Stage 3** | **Stage 4** |
| No. events | 27 | 60 | 1,143 | 256 | 1,071 |
| No. participants | 5,174 | 7,544 | 20,838 | 11,243 | 5,825 |
| Crude | Referent | 1.71 [1.08, 2.72] | 12.80 [8.68, 18.8] | 4.77 [3.18, 7.14] | 57.13 [38.7, 84.3] |
| Multivariable model 1[a] | Referent | 1.13 [0.71, 1.79] | 2.10 [1.42, 3.11] | 2.32 [1.54, 3.48] | 5.28 [3.56, 7.84] |
| Multivariable model 2[b] | Referent | 1.11 [0.70, 1.77] | 1.79 [1.20, 2.67] | 2.13 [1.40, 3.24] | 4.39 [2.93, 6.58] |
| Multivariable model 3[c] | Referent | 1.11 [0.70, 1.77] | 1.78 [1.19, 2.65] | 2.13 [1.40, 3.23] | 4.33 [2.89, 6.49] |
| **Unweighted Models to Contextualize the Unweighted Adjusted Cumulative Incidence Function Estimates*** | | | | | |
| | **Stage 0** | **Stage 1** | **Stage 2** | **Stage 3** | **Stage 4** |
| Crude | Referent | 1.70 [1.07, 2.69] | 11.70 [7.94, 17.25] | 4.63 [3.09, 6.92] | 43.31 [29.3, 63.8] |
| Multivariable model 1[a] | Referent | 1.23 [0.78, 1.94] | 2.47 [1.68, 3.64] | 2.44 [1.63, 3.64] | 5.38 [3.63, 7.95] |
| Multivariable model 2[b] | Referent | 1.17 [0.74, 1.84] | 1.98 [1.33, 2.95] | 1.98 [1.31, 2.99] | 4.16 [2.78, 6.22] |
| Multivariable model 3[c] | Referent | 1.16 [0.74, 1.84] | 1.97 [1.33, 2.94] | 1.98 [1.31, 2.99] | 4.14 [2.77, 6.19] |

[a] Multivariable model 1: age, sex, race or ethnicity, smoking status, survey year.

[b] Multivariable model 2: adjusted for Model 2 + BMI, waist circumference, hemoglobin A1c, systolic blood pressure, total cholesterol, statin use, ACE/ARB use, UACR, eGFR.

[c] Multivariable model 3: adjusted for Model 3 + history of chronic respiratory disease, history of liver disease, history of cancer.

* These models incorporate non-cardiovascular mortality as competing events.

| Supplemental Table 12. Adjusted Cumulative Incidence and Absolute Risk Differences for All-Cause Mortality | | |
|---|---|---|
| **Adjusted cumulative incidence, % (95% CI)** | **10-year** | **15-year** |
| Stage 0 | 12.2 (10.8-13.6) | 19.9 (18-21.6) |
| Stage 1 | 11.2 (10.2-12.2) | 18.6 (17.1-20.0) |
| Stage 2 | 12.9 (12.5-13.3) | 20.9 (20.2-21.5) |
| Stage 3 | 15.8 (15.0-16.7) | 24.7 (23.6-25.8) |
| Stage 4 | 19.4 (18.7-20.0) | 29.1 (28.2-30.0) |
| **Absolute risk difference, % (95% CI)** | **10-year** | **15-year** |
| CKM Stage 4 - Stage 0 | 7.2 (5.6-8.7) | 9.2 (7.0-11.3) |
| CKM Stage 4 - Stage 1 | 8.1 (6.9-9.3) | 10.5 (8.8-12.2) |
| CKM Stage 4 - Stage 2 | 6.4 (5.7-7.2) | 8.2 (7.1-9.3) |
| CKM Stage 4 - Stage 3 | 3.5 (2.5-4.6) | 4.4 (2.9-5.8) |

| Supplemental Table 13. Association between CKM syndrome stage and all-cause mortality. | | | | | |
|---|---|---|---|---|---|
| **Models Incorporating Survey Weights** | | | | | |
| | **Stage 0** | **Stage 1** | **Stage 2** | **Stage 3** | **Stage 4** |
| No. events | 207 | 347 | 3,942 | 1,027 | 2,646 |
| No. participants | 5,174 | 7,544 | 20,838 | 11,243 | 5,825 |
| Crude | Referent | 1.27 [1.06, 1.52] | 5.34 [4.60, 6.20] | 2.23 [1.87, 2.65] | 19.48 [16.75, 22.66] |
| Multivariable model 1[a] | Referent | 0.88 [0.73, 1.07] | 1.13 [0.96, 1.33] | 1.34 [1.11, 1.61] | 2.19 [1.82, 2.63] |
| Multivariable model 2[b] | Referent | 0.96 [0.79, 1.16] | 1.13 [0.94, 1.35] | 1.45 [1.19, 1.77] | 2.12 [1.75, 2.57] |
| Multivariable model 3[c] | Referent | 0.98 [0.81, 1.19] | 1.14 [0.96, 1.36] | 1.46 [1.20, 1.77] | 2.06 [1.72, 2.48] |
| **Unweighted Models to Contextualize the Unweighted Adjusted Cumulative Incidence Estimates** | | | | | |
| | **Stage 0** | **Stage 1** | **Stage 2** | **Stage 3** | **Stage 4** |
| Crude | Referent | 1.24 [1.04, 1.48] | 5.59 [4.86, 6.43] | 2.42 [2.08, 2.81] | 17.80 [15.40, 20.50] |
| Multivariable model 1[a] | Referent | 0.86 [0.72, 1.02] | 1.07 [0.93, 1.24] | 1.27[1.09, 1.48] | 1.98 [1.71, 2.30] |
| Multivariable model 2[b] | Referent | 0.89 [0.74, 1.06] | 1.07 [0.92, 1.25] | 1.42 [1.21, 1.67] | 1.97 [1.69, 2.30] |
| Multivariable model 3[c] | Referent | 0.90 [0.75, 1.07] | 1.07 [0.92, 1.25] | 1.41 [1.20, 1.66] | 1.90 [1.63, 2.22] |

[a] Multivariable model 1: age, sex, race or ethnicity, smoking status, survey year.
[b] Multivariable model 2: adjusted for Model 2 + BMI, waist circumference, hemoglobin A1c, systolic blood pressure, total cholesterol, statin use, ACE/ARB use, UACR, eGFR.
[c] Multivariable model 3: adjusted for Model 3 + history of chronic respiratory disease, history of liver disease, history of cancer.

| Supplemental Table 14. Association between CKM syndrome stage and non-cardiovascular mortality | | | | | |
|---|---|---|---|---|---|
| | **Stage 0** | **Stage 1** | **Stage 2** | **Stage 3** | **Stage 4** |
| No. events | 180 | 287 | 2,799 | 771 | 1,575 |
| No. participants | 5,174 | 7,544 | 20,838 | 11,243 | 5,825 |
| Crude | Referent | 1.24 [1.01, 1.52] | 4.67 [4.00, 5.50] | 2.00 [1.64, 2.44] | 15.61 [13.08, 18.61] |
| Multivariable model 1[a] | Referent | 0.87 [0.70, 1.08] | 1.02 [0.85, 1.22] | 1.22 [1.00, 1.50] | 1.87 [1.51, 2.30] |
| Multivariable model 2[b] | Referent | 0.96 [0.77, 1.20] | 1.07 [0.88, 1.29] | 1.45 [1.17, 1.81] | 1.94 [1.56, 2.41] |
| Multivariable model 3[c] | Referent | 0.98 [0.79, 1.22] | 1.07 [0.89, 1.29] | 1.44 [1.16, 1.79] | 1.85 [1.50, 2.30] |

[a] Multivariable model 1: age, sex, race or ethnicity, smoking status, survey year.

[b] Multivariable model 2: adjusted for Model 2 + BMI, waist circumference, hemoglobin A1c, systolic blood pressure, total cholesterol, statin use, ACE/ARB use, UACR, eGFR.

[c] Multivariable model 3: adjusted for Model 3 + history of chronic respiratory disease, history of liver disease, history of cancer.

| **Supplemental Table 15. Limitations to defining CKM syndrome in NHANES.** | |
|---|---|
| Hyperlipidemia | Low-density lipoprotein cholesterol and triglycerides are missing in approximately 50% of NHANES participants, as they were required to perform the labs in a fasting state. |
| Metabolic syndrome | Due to missingness in fasting labs (triglycerides and blood glucose), we defined metabolic syndrome using the modified parameters as described in Table S1. The definition included hemoglobin A1c (rather than fasting blood glucose) and did not include elevated triglycerides. |
| Obesity in Asian participants | The CKM syndrome guidelines recommend alternative body mass index and waist circumference thresholds for individuals of Asian descent. NHANES began specifically reporting ‘Asian’ as a race category in 2011, preventing use of the Asian specific criteria across all survey years. For consistency, we did not apply Asian specific thresholds in our definition of CKM syndrome. |
| End-stage kidney disease and dialysis use | Dialysis use is asked in NHANES as “have you undergone dialysis in the last 12 months,” which could include individuals with either end-stage kidney disease requiring chronic renal replacement therapy or those who received dialysis for severe acute kidney injury (with potential for renal recovery). |
| Subclinical cardiovascular disease | NHANES does not include coronary artery calcification scores or echocardiography. Cardiac biomarkers (such as high-sensitivity troponin and NT-proBNP) were only collected in the 1999-2004 examinations. |
| Clinical cardiovascular disease | NHANES does not include self-reported peripheral artery disease or atrial fibrillation. Ankle-Brachial Index was only obtained in the 1999-2004 examinations. |
| PREVENT equation | The PREVENT equation is validated for predicting cardiovascular risk among individuals age ≥30 years. In this analysis, it was also applied to participants ages 20-29 years. |
